# An Exploratory Systematic Review and Meta-analysis on Period Poverty

**DOI:** 10.1101/2023.01.18.23284706

**Authors:** Gayathri Delanerolle, Xiaojie Yang, Heitor Cavalini, Om Kurmi, Camilla Rostivik, Ashish Shetty, Lucky Saraswat, Julie Taylor, Sana Sajid, Shanaya Rathod, Jian Qing Shi, Peter Phiri

## Abstract

**Background:** *Period poverty* is a global health and social issue that needs to be addressed. The primary aim of this study was to provide a comprehensive understanding on period poverty, including outcomes associated menstruation.

**Methods:** All observational and randomised clinical trials reporting menstruation challenges, menstrual poverty and menstrual products were included. Our search strategy included multiple electronic databases of PubMed, Web of Science, ScienceDirect, ProQuest and EMBASE. Studies published in a peer review journal in English between the 30^th^ of April 1980 and the 30^th^ of April 2022 were included. The Newcastle-Ottawa Scale (NOS) was used to assess the risk of bias (RoB) of the systematic included studies. Pooled odds ratios (ORs) together with 95% confidence intervals (CIs) are reported overall and for sub-groups.

**Results:** A total of 80 studies were systematically selected, where 38 were included in the meta-analysis. Of the 38 studies, 28 focused on children and young girls (i.e., 10-24 years old) and 10 included participants with a wider age range of 15-49 years. The prevalence of using disposable sanitary pads was 45% (95% CI = [0.35,0.58]). The prevalence of menstrual education pre-menarche was 68% (95% CI = [0.56, 0.82]). The prevalence of good MHM was 39% (95% CI = [0.25, 0.61]). Women in rural area (OR = 0.30, 95% CI = [0.13, 0.69]) were 0.70 times less likely to have good MHM practices than those living in urban area.

**Discussion:** There was a lack of evidence, especially from low- and middle-income countries. Further research to better understand the scope and prevalence of period poverty should be considered. This will enable the development of improved policies to increase access to menstrual products and medical support where necessary.

**Funding:** Not Applicable

**Registration:** A systematic methodology was developed and published (CRD42022339536).

## Introduction

The World Health Organisation (WHO) defines *health* as complete mental, physical, and social well-being, thus not a mere absence of a disease or infirmity. For women, menstrual health is integral to maintain their overall health as menstruation occurs between menarche and menopause, which may have a significant impact on their mental, physical, and social wellbeing. Menstruation, or periods, is a biological process that is part of nearly every biological female’s life and is defined as cyclical bleeding that occurs as a result of the regeneration of the uterine endometrium corpus. Clinically, the *normal* menstrual process of 4 phases across a cycle of 28-35 days.^1-2^ The regularity of these cycle, duration of each of the bleeding episodes within a cycle and the volume or heaviness of the bleed varies across women and can change throughout an individual’s lifespan. ^1-2^ All women do not experience *normal* menstrual bleeding, with approximately 30% experiencing alterations to their pattern or volume of menstrual flow due to multiple aetiologies. ^3-4^ Many women also report symptoms such as pain, anxiety, fatigue, dysmenorrhea, and depression associated with their menstrual cycle that may require clinical involvement to diagnose potential reproductive health issues such as premenstrual dysphoric disorder, premenstrual syndrome, or endometriosis. ^1-3^ To promote positive health and wellbeing outcomes to all genders and clinicians, it is important to understand menstrual cycles and menstrual health that can be promoted in the first instance by way of menstrual health literacy and various public interventions such as maintaining good hygiene practices and access to menstrual products.

Access to menstrual products is as vital as access to other hygiene products. The WHO and United Nations International Children’s Emergency Fund (UNICEF) has reported that many girls miss school and put their lives on hold to remain at home during their menstruation days due to a number of reasons, including access to toilets or menstrual products^5-6^. This is commonly reported as *period or menstruation poverty*.^7^ Period poverty is a global health issue impacting people who do not have access to hygienic and safe menstrual products. This is particularly important for regions with conflicts and disasters, which leave menstruating people with minimal or no access to safe menstrual products, clean and toilets. This could lead to the use of unconventional methods to manage the bleeding such as the use of clothing, rags or sitting on old tin cans. ^7-9^ Ancient traditions such as *Chhaupadi* practices could further risk girls and women from securely managing their menstruation.^10^ Chhaupadi is practiced in some far western rural regions in Nepal where young girls are banished into sheds or mud huts during menstruation or even longer as they believe this brings ill health or bad luck to the families. ^10^ Often these people have little or no access to washing facilities or supplies leading to health issues, including physical and psychological hardship. ^10-11^ Despite Chhaupadi being illegal in Nepal since 2005, Action Aid reports that it is practiced in some communities to date.^11^ Whilst poverty and stigma impact the right for a girl child’s education, especially in low-middle-income countries, The United Nations Educational, Scientific and Cultural Organisation (UNESCO) reports that 1 in 10 girls in Africa alone misses school during their menstruation. ^12-15^ Missing school could lead to dropping out, risking child marriage and pregnancy at a younger age, as reported by Action Aid^16^.

It has been reported for many decades that menstrual poverty is associated with stigma and shame and impacts the dignity and overall wellbeing. Despite being a developed country, over 37% of women in the UK have experienced periods shaming by way of isolation, bullying and jokes, based on an Action Aid survey report^17^. Approximately 40% of women reported being humiliated by their partners, while over half of UK women said they were embarrassed when they got their periods for the first time. In addition, over 52% reported they hide sanitary products when taking these to the toilet to prevent anyone else from being embarrassed, whilst 43% reported they felt people would make inappropriate remarks. The New York Post reported similar findings from a study commissioned by THINX, which indicated 58% of women felt embarrassed during the menstruation period whilst 42% experienced period-shaming, where 1 in 5 of those women reported these feelings were due to comments made by male friends. ^18^

It is evident that period poverty appears to be a global phenomenon, and key sociological as well as clinical features may differ due to varying risk factors in diverse geographical regions. To identify the impact of period poverty in diverse populations and common denominators observed between low-middle-income countries (LMICs) and high-income countries, it is vital to better understand current gaps in knowledge, policies and practice. To achieve this, we developed the PLATO project with the first component focusing on an evidence synthesis of the existing peer reviewed literature.

## Methods

A systematic methodology was developed and published as a protocol in PROSPERO (CRD42022339536) to explore period poverty. A meta-analysis was conducted in addition to two key thematic variables identified through the systematic review of homelessness, infections, lived experiences and mental health impact due to menstruation.

Within the context of this study, rural and urban areas of the study were defined by natural administrative division of the location as reported within the peer review publications. The division of LMIC, middle-low-income countries (MICs) and high-income countries (HICs) were defined based on the dividing standards of the World Banking Group.

The primary aim of this study was to provide a comprehensive understanding on period poverty, including outcomes associated menstruation such as affordability of menstrual products such as disposable sanitary pads, accessibility to menstruation education tools, adequate menstrual hygiene management (MHM) practice and urinary tract infections. The difference in MHM practices in a variety of contexts such as age groups, religious beliefs, parents’ educational status, and school absenteeism due to dysmenorrhea were also explored.

### Inclusion/Exclusion

All observational and randomised clinical trials reporting menstruation challenges, menstrual poverty and menstrual products were included. Studies published in a peer review journal in English between the 30^th^ of April 1980 and the 30^th^ of April 2022 were included. All editorials, letters to editors and commentaries, and papers published in languages other than English were excluded.

### Patient and Public Involvement

All the data used in this systematic review is publicly available. No further patient or public involvement was implemented for this paper.

### Search strategy

Our search strategy included multiple electronic databases of PubMed, Web of Science, ScienceDirect, ProQuest and EMBASE. Subject index terms used were: Menstrual education, anthropology, period poverty, pads, sanitary pads, sanitary facilities, menstrual hygiene, urinary tract infections, menstrual health, and women’s periods. The title and abstract of each publication were screened independently by two investigators. A consensus was reached for studies that were unsuitable for inclusion. Articles that were included were reviewed in in full independently by two investigators. These were re-reviewed independently prior to the data extraction. Difference of opinions and queries were resolved by the by the Principal Investigator and Chief Investigator.

### Data extraction

We developed an extraction template specific to the objectives of the study although the aim was to gather a wider dataset to ensure vital data was not missed to answer the research aims comprehensively.

Participants included in the study populations were those who live and/or are at risk of menstrual poverty. All studies reporting a menstrual product and/or an educational intervention associated with menstruation were extracted by way of the instruments, measures of tool and questionnaires. The final dataset was independently reviewed before the analysis commenced.

Participants included in the study populations were those who have experienced or are at risk of menstrual poverty. All studies reporting a menstrual product and/or an educational intervention associated with menstruation were extracted by way of the instruments, measures of tool and questionnaires. The final dataset was independently reviewed before the analysis commenced.

### Risk of bias

The Newcastle-Ottawa Scale (NOS) was used to asses the risk of bias (RoB) of the systematic included studies. A risk of bias table has been made available as a supplementary file. The RoB table reflects a fixed set of biases linked to the study design, conduct and reporting.

### Meta-analyses

Out of the 1432 studies screened, 1182 were excluded. Of the 250 studies assessed for eligibility, 170 were excluded. Hence, 80 studies were systematically included, and 38 were included in the meta-analysis (see Figure 1). The 38 studies were explored to obtain several indicators of period poverty, such as access to menstrual education tools, use of menstrual pads and MHM practice, as well as their related issues such as urinary tract infections, religious status, educational level of parents, geographical location including urban and rural areas, and the presence of a financial allowance.

**Figure 1.**
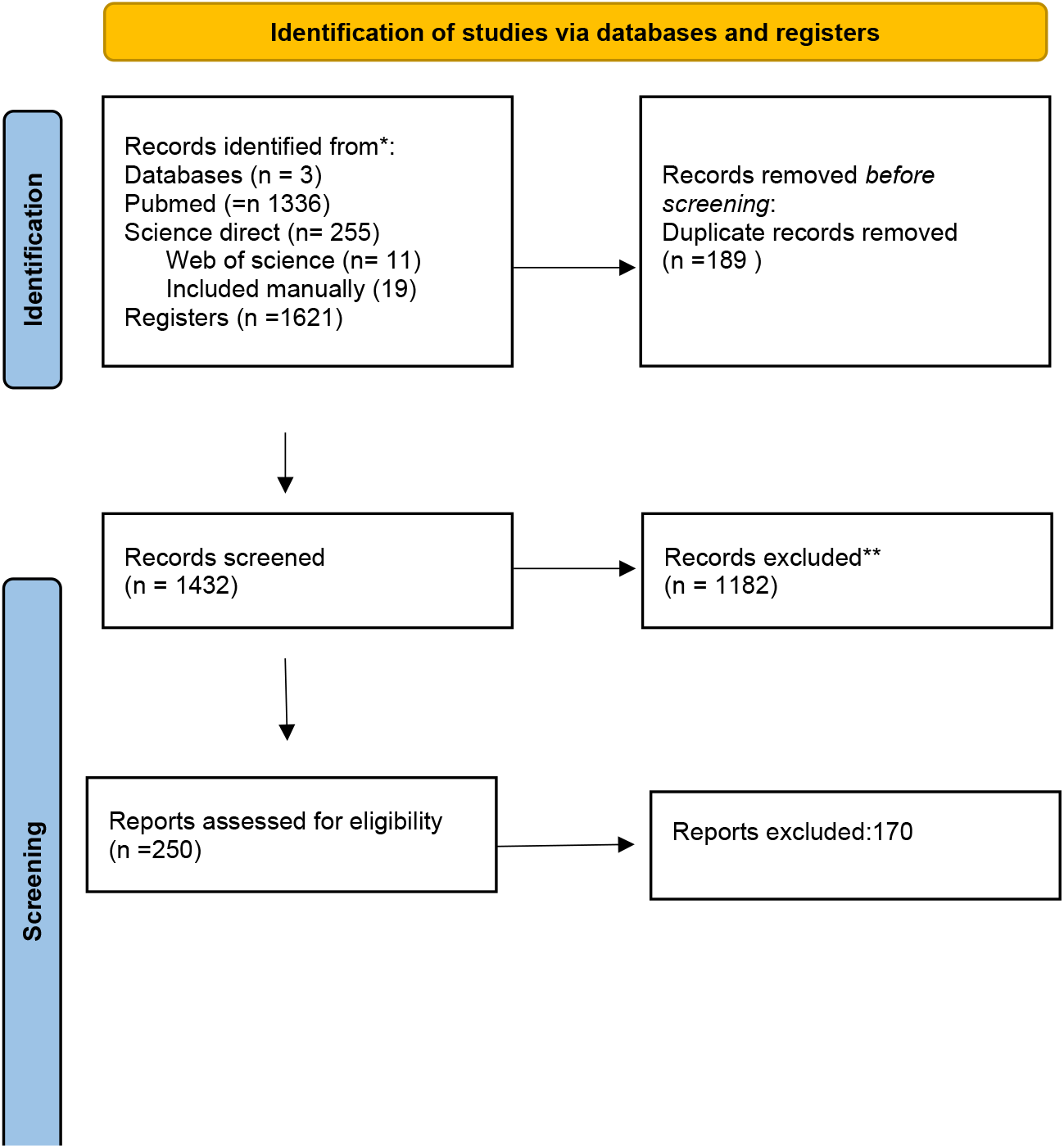
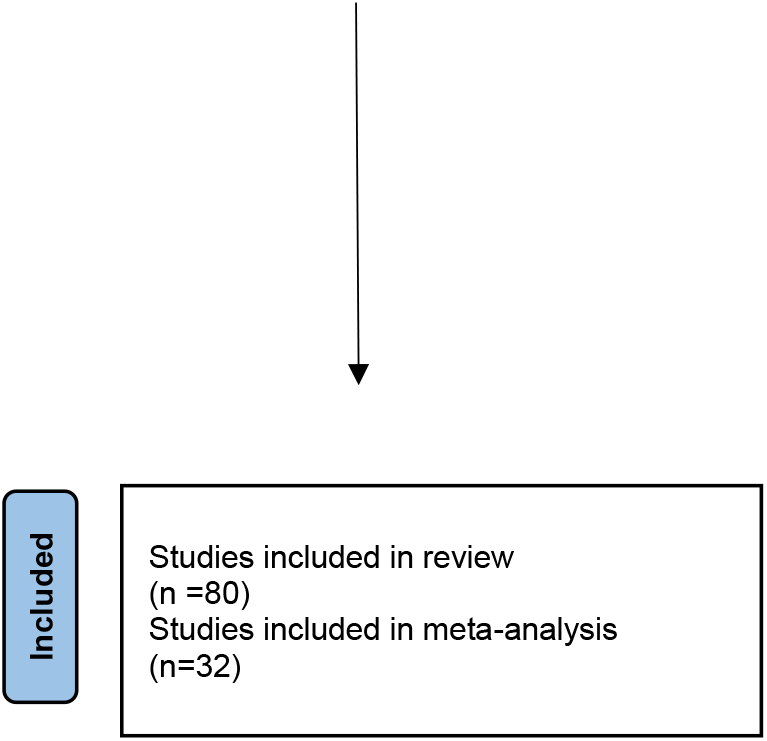
PRISMA 2020 Flow Diagram showing study selection.

To calculate the summary effect size across studies, meta-analysis of single proportions was applied to (a)-(c), and meta-analysis for comparison of two interventions was applied to (d)-(k). ^19-20^ Since almost all outcomes of interest in the current analysis were dichotomous, meta-analysis with binary data was performed, and accordingly the pooled odds ratio (OR) with a 95% confidence interval (CI) was used to access the effect of two interventions. ^20-21^ Statistical heterogeneity was evaluated by the commonly used measure *I*^2^ with p-value, and further *I*^2^ larger than 50% with a much small p-value indicates strong heterogeneity. In comparison, *I*^2^ less than 50% with a large p-value indicates fairly weak heterogeneity. ^22^ In the presence of high heterogeneity, the random effects model was employed; instead, the fixed effects model was used if there was weak or no heterogeneity. ^23^ In some cases, subgroup analysis was carried out to identify the sources of heterogeneity, and sensitivity analysis was conducted for mainly assessing robustness of the synthesized results. Finally, publication bias was addressed seriously in the discussion part. All statistical outputs were reported using R. ^24-25^

A systematic analysis was used for studies that were excluded from the meta-analysis including those reporting lived experiences and the mental health impact associated with menstruation.

## Results

Studies with limited discussion about menstrual products, menstruation knowledge and menstrual hygiene management (MHM) practice were excluded, resulting in a final dataset of 80 studies (see Table 2). Of the 80 studies, 38 studies were selected for meta-analysis. Of 38 the studies, 34 were from lower middle-income countries (LMICs) and 4 from developed countries (non-LMICs).

**Table 1.**
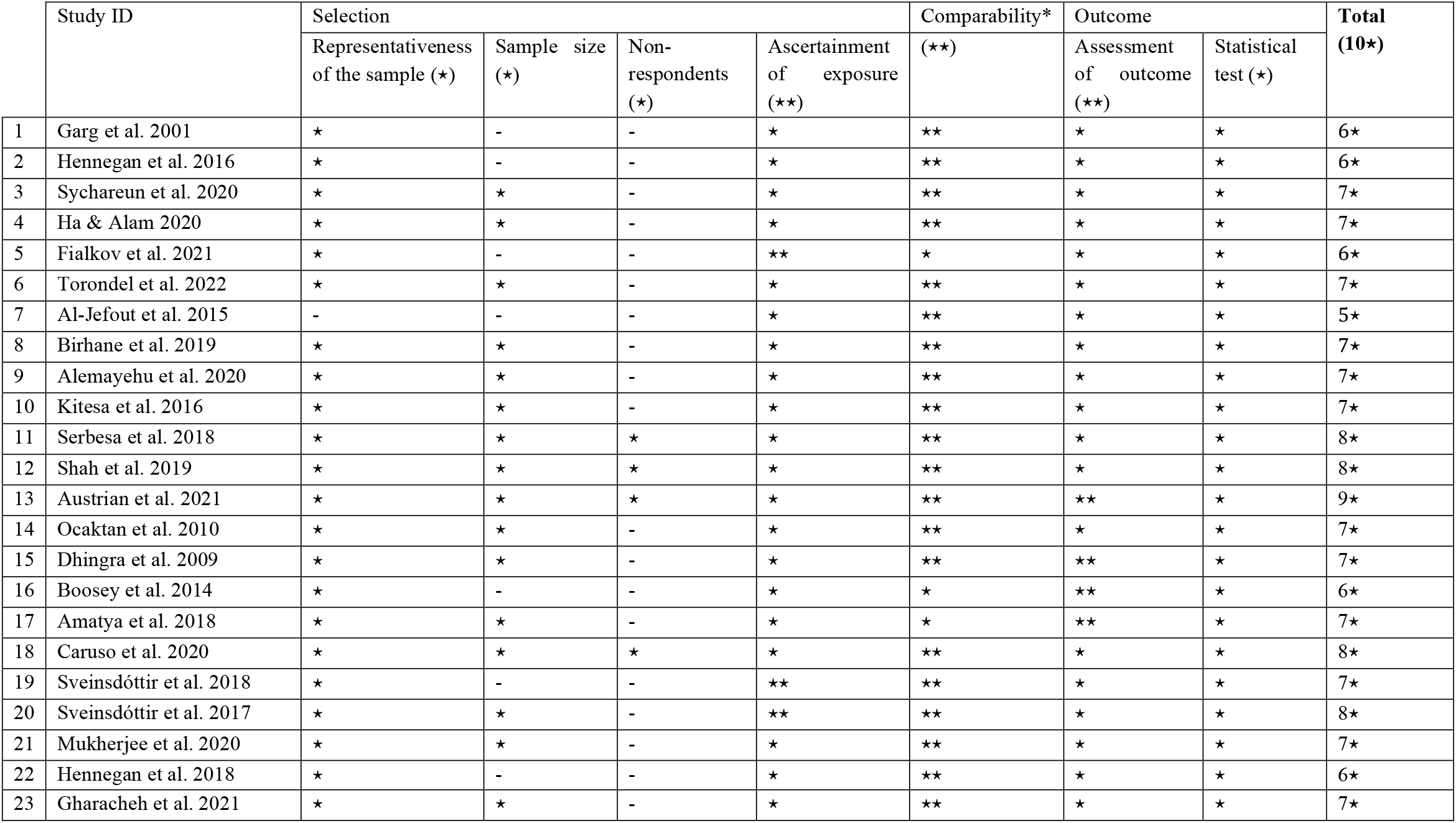
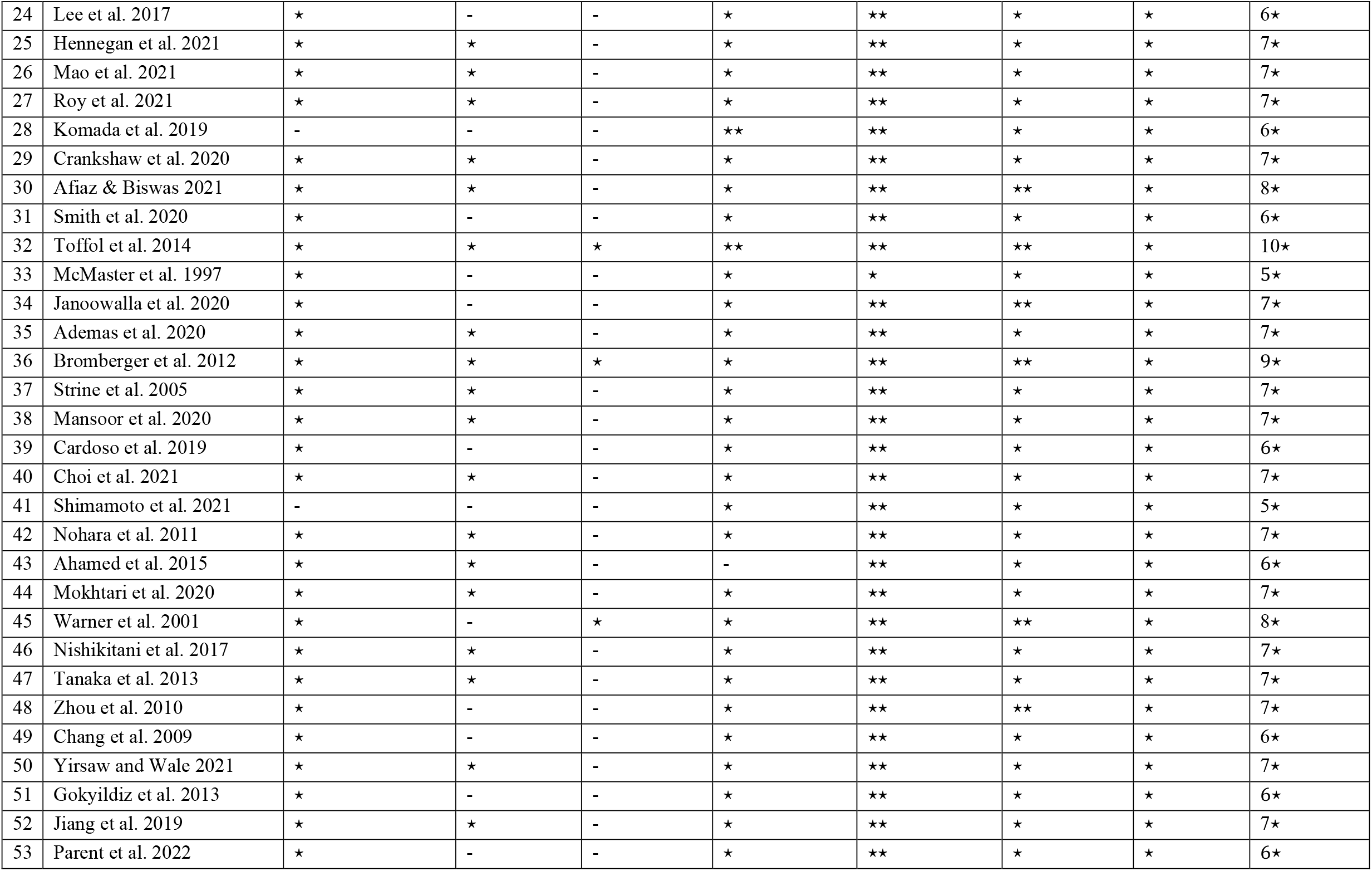
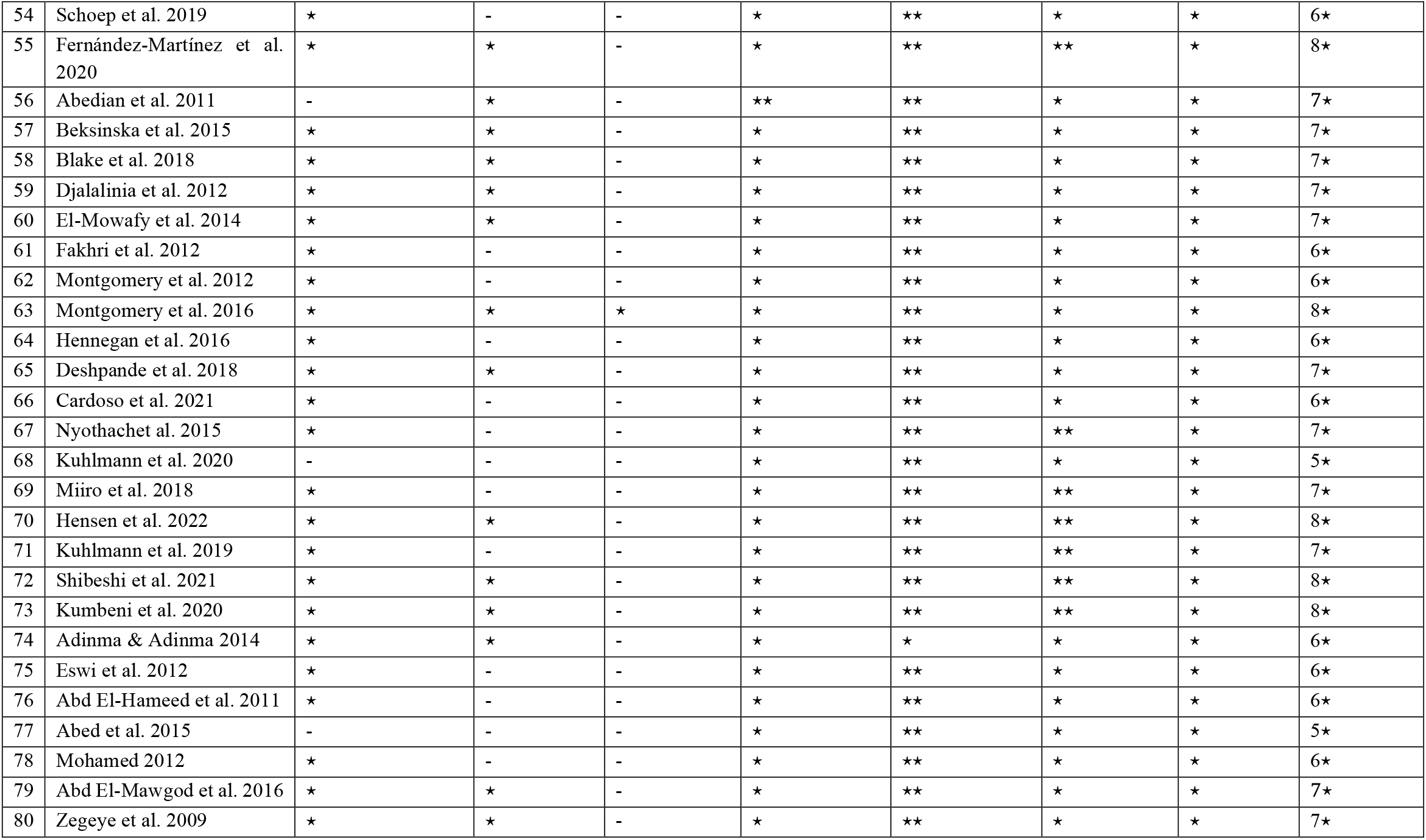
Quality Assessment of studies using a modified Newcastle-Ottowa scale (Modesti et al. 2016)

**Table 2.** Characteristics of the Studies included in the Systematic Review.

| Study ID | Authors | Year | Study Type | Sample Size | Country | Mean Age | Meta-analysis inclusion Y/N |
| --- | --- | --- | --- | --- | --- | --- | --- |
| 1 | Garg et al. | 2001 | Epidemiological and sociological study | 380 | India |  | Y |
| 2 | Hennegan et al. | 2016 | Cross-sectional study | 201 | Uganda | 14.2 | Y |
| 3 | Sychareun et al. | 2020 | Cross-sectional study | 343 | LAO | 15.6 | Y |
| 4 | Ha & Alam | 2020 | Cross-sectional study design with systematic random sampling | 589 | Bangladesh | 15.5 | Y |
| 5 | Fialkov et al. | 2021 | Pre- and post-test design that compared six cohort groups. | 311 | Kenya |  | N |
| 6 | Torondel et al. | 2022 | Nested within a pair-matched cohort study | 1045 | India | 27 | Y |
| 7 | Al-Jefout et al. | 2015 | Cross-sectional study | 272 | Jordanian | 22 | Y |
| 8 | Birhane et al. | 2019 | Cross-sectional study | 466 | Ethiopia | 15.5 | Y |
| 9 | Alemayehu et al. | 2020 | Cross-sectional study | 301 | Ethiopia | 15.87 | Y |
| 10 | Kitesa et al. | 2016 | Cross-sectional study | 430 | Ethiopia | 16 | Y |
| 11 | Serbesa et al. | 2018 | Cross-sectional study | 310 | Ethiopia | 15.72 | Y |
| 12 | Shah et al. | 2019 | Cross-sectional study | 331 | Gambia | 15.3 | Y |
| 13 | Austrian et al. | 2021 | Cluster RCT | 3489 | Kenya | 14.8 | N |
| 14 | Ocaktan et al. | 2010 | Cross-sectional study | 400 | Turkey | 32.19 | Y |
| 15 | Dhingra et al. | 2009 | Cross-sectional study | 200 | India | 13.97 | Y |
| 16 | Boosey et al. | 2014 | Cross-sectional study | 140 | Uganda | 14.45 | N |
| 17 | Amatya et al. | 2018 | Cross-sectional mixed-methods study | 104 | Nepal | 15 | N |
| 18 | Caruso et al. | 2020 | Cross-sectional study | 878 | India | 26.8 | Y |
| 19 | Sveinsdóttir et al. | 2018 | Cross-sectional study | 319 | Iceland | 30 | Y |
| 20 | Sveinsdóttir et al. | 2017 | Cross-sectional study | 319 | Iceland | 30 | N |
| 21 | Mukherjee et al. | 2020 | Cross-sectional study | 1342 | Nepal |  | N |
| 22 | Hennegan et al. | 2018 | Cross-sectional study | 2934 | Nigeria | 26.66 | Y |
| 23 | Gharacheh et al. | 2021 | Cross-sectional study | 515 | Iran | 29.61 | N |
| 24 | Lee et al. | 2017 | Prospective observational cohort study | 1495 | USA | 46.8 | N |
| 25 | Hennegan et al. | 2021 | Secondary data analysis | 11806 | Burkina Faso, Niger, Nigeria |  | N |
| 26 | Mao et al. | 2021 | Cross-sectional study | 156055 | China | 26.32 | N |
| 27 | Roy et al. | 2021 | Secondary data analysis | 94034 | India |  | Y |
| 28 | Komada et al. | 2019 | Cross-sectional study | 150 | Japan | 18.8 | N |
| 29 | Crankshaw et al. | 2020 | Mixed-method study | 472 | South Africa | 17.5 | Y |
| 30 | Afiaz & Biswas | 2021 | Cross-sectional study | 54242 | Bangladesh | 29 | Y |
| 31 | Smith et al. | 2020 | Secondary data analysis | 38257 | Uganda, Kenya, Ethiopia etc. |  | Y |
| 32 | Toffol et al. | 2014 | Cross-sectional study | 4391 | Finland | 56.2 | N |
| 33 | McMaster et al. | 1997 | exploratory phase of the study | 50 | Zimbabwe |  | N |
| 34 | Janoowalla et al. | 2020 | Prospective cohort study | 240 | Rwanda | 19.1 | Y |
| 35 | Ademas et al. | 2020 | Cross-sectional study | 602 | Ethiopia |  | Y |
| 36 | Bromberger et al. | 2012 | Multisite study | 934 | USA |  | N |
| 37 | Strine et al. | 2005 | Cross-sectional study | 11648 | USA |  | N |
| 38 | Mansoor et al. | 2020 | Cross-sectional study | 1777 | Pakistan | 20.38 | Y |
| 39 | Cardoso et al. | 2019 | Baseline data from a larger RCT | 1800 | Nepal | 34.5 | N |
| 40 | Choi et al. | 2021 | Cross-sectional study | 8658 | Korea | 35.1 | Y |
| 41 | Shimamoto et al. | 2021 | Self-reporting questionnaire survey | 6048 | Japan |  | N |
| 42 | Nohara et al. | 2011 | Cross-sectional study | 2166 | Japan |  | N |
| 43 | Ahamed et al. | 2015 | Cross-sectional study | 344 | India | 28 | Y |
| 44 | Mokhtari et al. | 2020 | Cross-sectional study | 164 | Iran | 27.78 | N |
| 45 | Warner et al. | 2001 | Cross-sectional study | 952 | Scotland |  | N |
| 46 | Nishikitani et al. | 2017 | Cross-sectional study | 505 | Japan |  | N |
| 47 | Tanaka et al. | 2013 | Online survey | 19254 | Japan | 33.6 | N |
| 48 | Zhou et al. | 2010 | Cross-sectional study | 1642 | China | 37 | N |
| 49 | Chang et al. | 2009 | Cross-sectional survey | 1095 | Taiwan |  | N |
| 50 | Yirsaw and Wale | 2021 | Cross-sectional study | 713 | Ethiopia | 21.13 | N |
| 51 | Gokyildiz et al. | 2013 | Case-control study | 295 | Turkey |  | N |
| 52 | Jiang et al. | 2019 | Cross-sectional study | 12881 | China |  | N |
| 53 | Parent et al. | 2022 | Cross-sectional study | 1153 | France | 31.7 | Y |
| 54 | Schoep et al. | 2019 | Cross-sectional study | 42879 | Netherlands | 28.7 | N |
| 55 | Fernández-Martínez et al. | 2020 | Cross-sectional study | 7208 | Spain | 19.51 | N |
| 56 | Abedian et al. | 2011 | RCT | 165 | Iran |  | N |
| 57 | Beksinska et al. | 2015 | Randomized two-period Cross-over trial | 124 | South Africa | 29 | N |
| 58 | Blake et al. | 2018 | Mixed-methods evaluation | 636 | Ethiopia | 13.45 | Y |
| 59 | Djalalinia et al. | 2012 | Community-based participatory research | 1823 | Iran |  | N |
| 60 | El-Mowafy et al. | 2014 | Quasi-experimental study | 234 | Egypt |  | N |
| 61 | Fakhri et al. | 2012 | Quasi-experimental study | 698 | Iran | 15.7 | N |
| 62 | Montgomery et al. | 2012 | Non-randomized trial | 120 | Ghana | 15.7 | N |
| 63 | Montgomery et al. | 2016 | Cluster quasi-randomised controlled trial | 1124 | Uganda |  | N |
| 64 | Hennegan et al. | 2016 | Secondary data analysis | 205 | Uganda. | 14.2 | Y |
| 65 | Deshpande et al. | 2018 | Cross-sectional study | 100 | India |  | Y |
| 66 | Cardoso et al. | 2021 | Online survey | 471 | United States | 20.6 | N |
| 67 | Nyothachet al. | 2015 | Retrospective study |  | Kenya |  | N |
| 68 | Kuhlmann et al. | 2020 | Cross-sectional study | 58 | USA | 15.21 | N |
| 69 | Miiró et al. | 2018 | Cross-sectional study | 352 | Uganda | 15.6 | Y |
| 70 | Hensen et al. | 2022 | Mixed-methods analysis | 7546 | Zambia. |  | N |
| 71 | Kuhlmann et al. | 2019 | Cross-sectional study | 183 | USA | 35.8 | Y |
| 72 | Shibeshi et al. | 2021 | Cross-sectional study | 1078 | Ethiopia | 17.35 | Y |
| 73 | Kumbeni et al. | 2020 | Cross-sectional study | 705 | Ghana |  | Y |
| 74 | Adinma & Adinma | 2014 | Cross-sectional study | 550 | Nigeria |  | Y |
| 75 | Eswi et al. | 2012 | Cross-sectional study | 200 | Egypt | 15.45 | N |
| 76 | Abd El-Hameed et al. | 2011 | Cross-sectional study | 160 | Egypt | 17.2 | N |
| 77 | Abed et al. | 2015 | Cross-sectional study | 100 | Egypt | 14.25 | Y |
| 78 | Mohamed | 2012 | Cross-sectional study | 885 | Egypt | 16 | Y |
| 79 | Abd El-Mawgod et al. | 2016 | Cross-sectional study | 344 | Saudi Arabia | 16.2 | Y |
| 80 | Zegeye et al. | 2009 | Cross-sectional study | 612 | Ethiopia | 16.9 | Y |

### Meta-Analysis

#### Prevalence of Using Disposable Sanitary Pads

We explored the link of disposable sanitary pads as an indicator of period poverty. A meta-analysis of single proportions was applied to 32 studies with a sample of 212,459 women, that indicated a prevalence of 45% (95% CI = [0.35,0.58]). Figure 2a shows the forest plot for 32 studies. The value of 100% of *I*^2^ (p-value = 0) indicates a significant statistical heterogeneity.

**Figure 2a.**
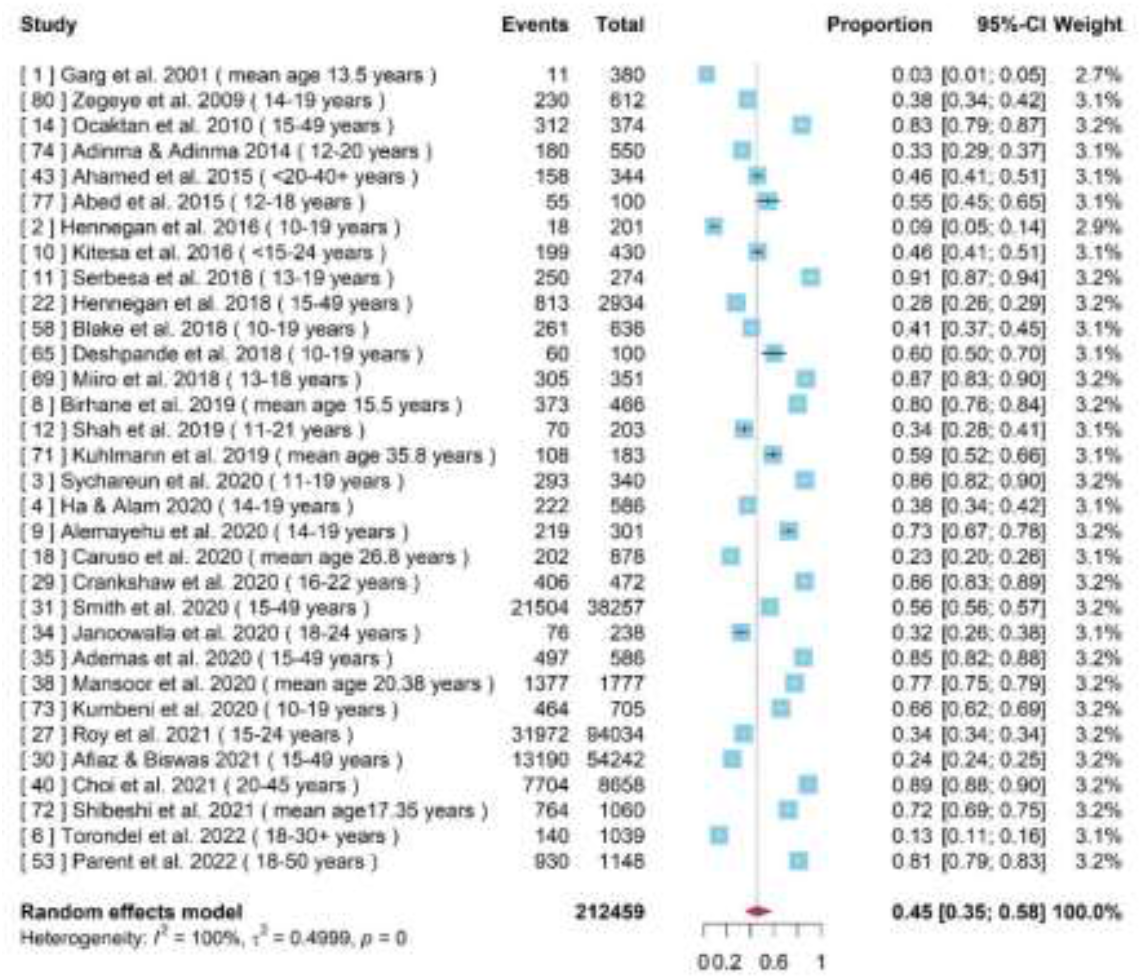
Forest plot shows the prevalence of using disposable sanitary pads across 32 studies.

To explore the sources of heterogeneity, a subgroup analysis was conducted using the geographical locations of the studies and demonstrated in a forest plot (Figure 2b). A statistically significant difference (p-value < 0.01) was identified between LMICs and non-LMICs using sanitary pads where the pooled prevalence was 43% (95% CI = [0.33, 0.56]) and 76% (95% CI = [0.60, 0.96]), respectively. Figure 2b also showed that heterogeneity remained unchanged in LMICs (*I*^2^ = 100%, p-value = 0) and non-LMICs (*I*^2^ = 98%, p-value < 0.01), indicating that the identified heterogeneity was not geographical location influenced.

**Figure 2b.**
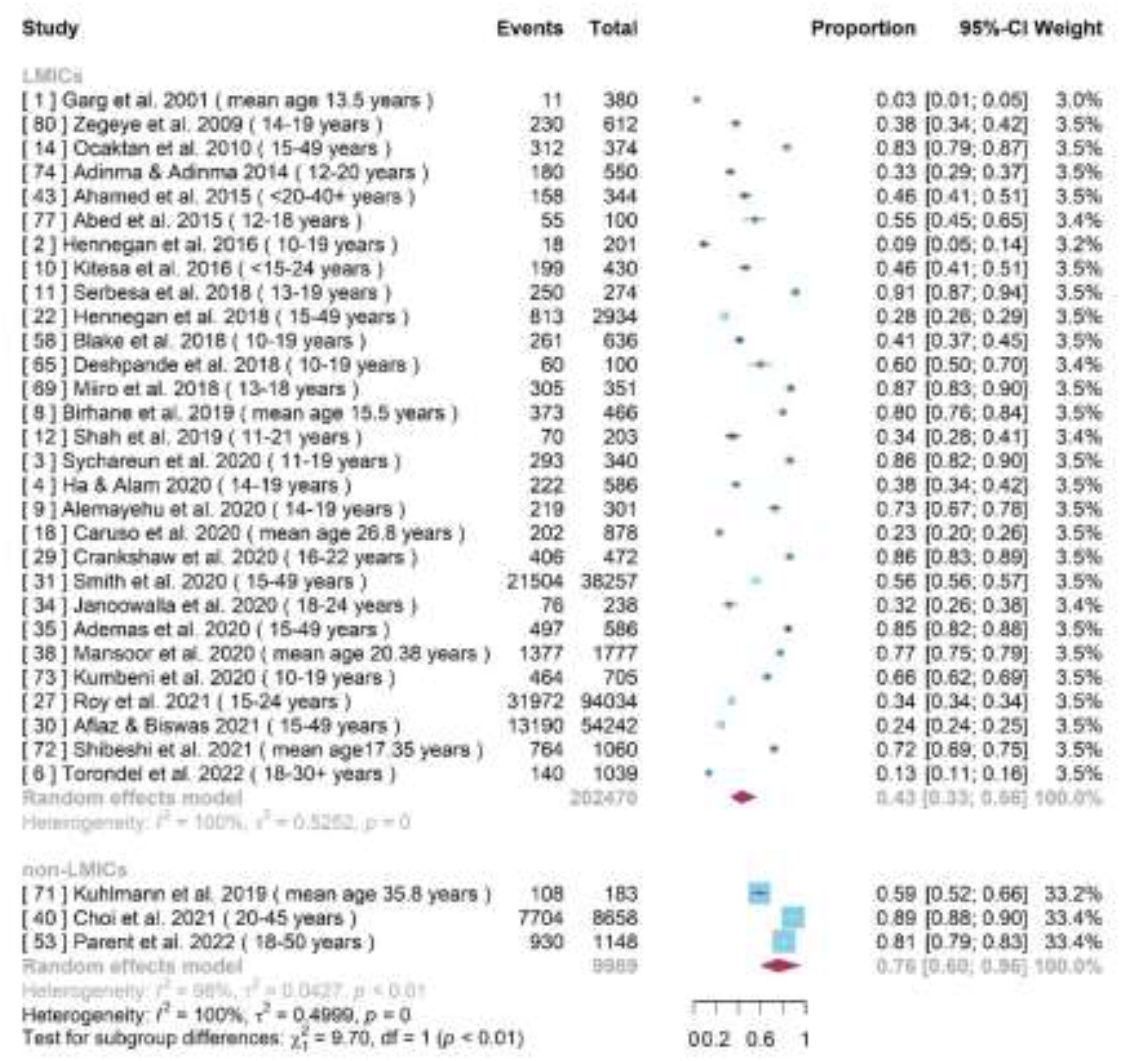
Forest plot shows the prevalence of using disposable sanitary pads in LMICs and non-LMICs, respectively.

#### Prevalence of Having Knowledge/Awareness on Menstruation before Menarche

Several surveys were meta-analysed to better understand adolescent girls’ menstrual education and pre-menarche awareness. Common survey questions notably included, “*were you familiar with menstruation before you got your first period* ^3^”, “*Information availability before reaching me*narche ^4^”, “*heard about menstruation before menarche* ^9^”, and “*prior knowledge about menstruation before menarche* ^11^” and “*awareness about menarche before its onset* ^79^”. This information was used to conduct a meta-analysis of 11 studies with a sample size of 4944 young women. A high heterogeneity was detected with *I*^2^ = 98% and p-value < 0.01) (Figure 3a). The random effects model reported the overall prevalence to be 68% (95% CI = [0.56, 0.82]).

**Figure 3a.**
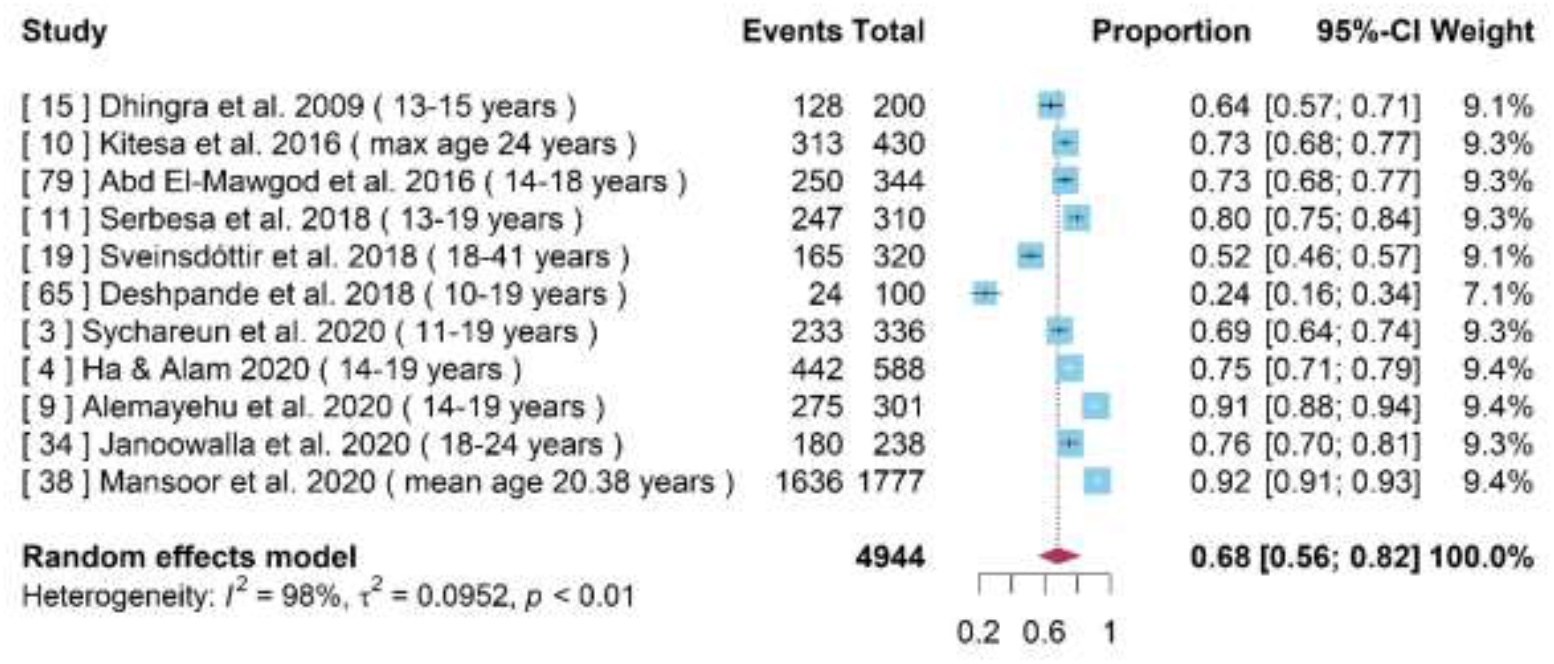
Forest plot for the prevalence of having knowledge/awareness on menstruation

#### Prevalence of Good Menstrual Hygiene Management (MHM) Practice

Good menstrual hygiene management (MHM) practice during menstruation is essential to prevent various other health issues such as urinary tract infections (UTIs). ^35^ MHM practice lacks a standardised definition although a consensus is that it is expected that throughout the bleeding phase, people require clean absorbents, adequate frequency of absorbent change, washing the body with soap and water, adequate disposal, and privacy for managing menstruation. All involved studies predesigned some practice-related questions in research studies to determine the level of MHM practice, defined simply as good or bad. Figure 3b demonstrates a forest plot of the prevalence of good MHM practice across ten studies with a total of 5432 women. The random effects model was used due to strong heterogeneity indicated by *I*^2^ = 99% and p-value < 0.01. The overall prevalence of good MHM practice was 39% (95% CI = [0.25, 0.61]).

**Figure 3b.**
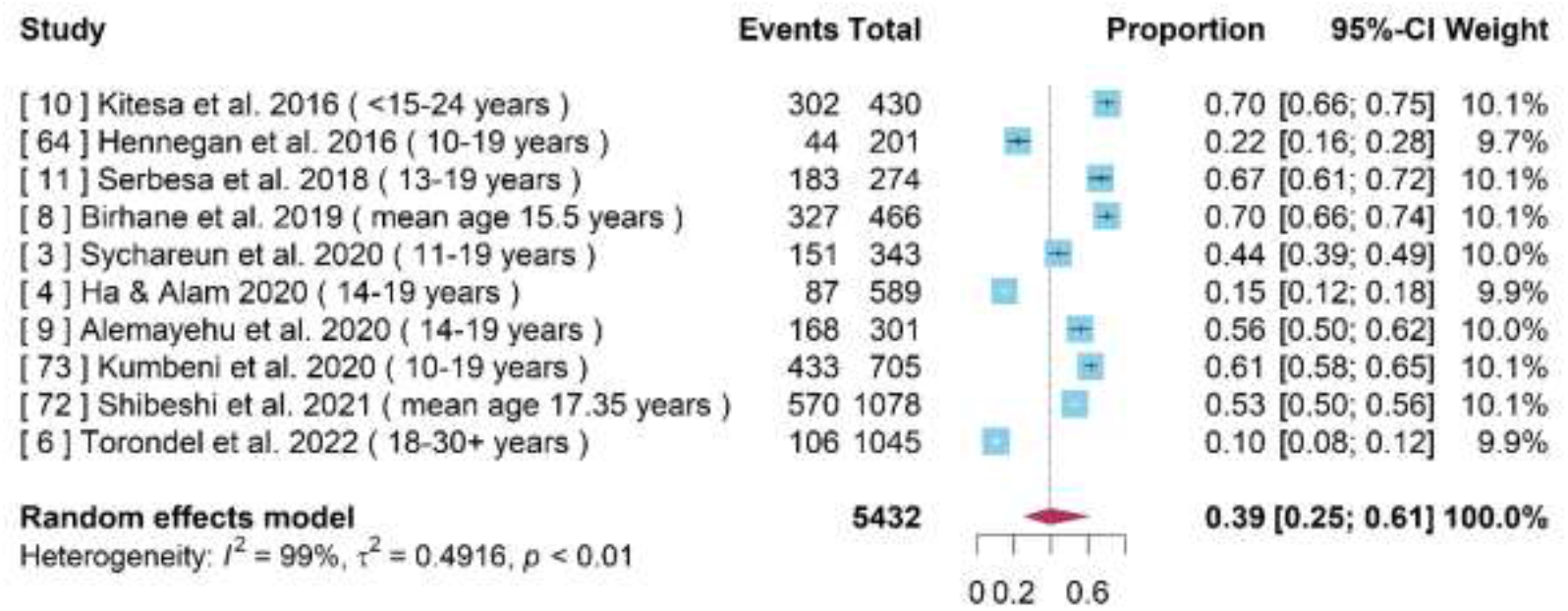
Forest plot for the prevalence of good MHM practice across ten studies.

#### Rural-urban Difference in MHM Practice Level (Good/Bad)

A total of 5 studies with a sample size of 2705 women reported differences of MHM practice levels within rural and urban settings. The pooled odds ratio (OR) of good MHM practice between rural and urban areas was 0.30 (95% CI = [0.13, 0.69]), indicating that women living in rural area were 0.70 times less likely to have good MHM practices in comparison to those living in an urban area. A high heterogeneity of 91% of *I*^2^ (p-value < 0.01) was identified (Figure 3c), possibly due to the differences in covariates, assessment tools and other factors.

**Figure 3c.**
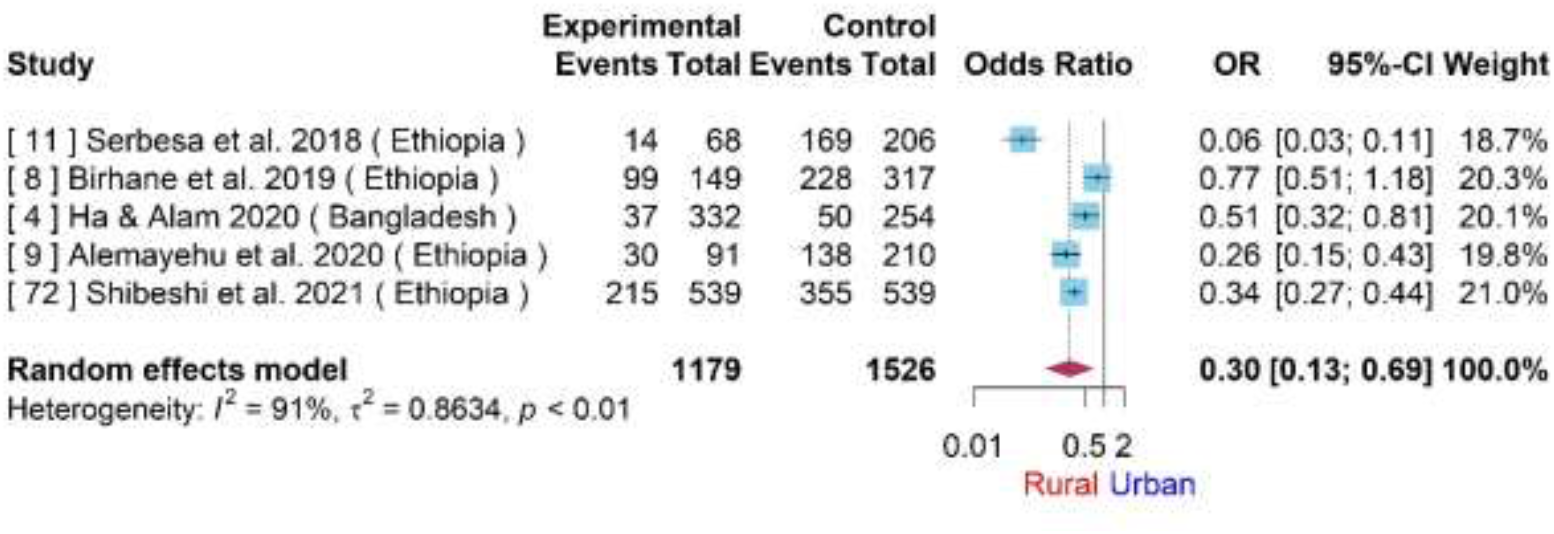
Forest plot for the rural-urban difference of MHM practice level (good/bad).

#### Difference of MHM Practice Level (Good/Bad) between Two Age Groups

Based on available data from 3 studies with a total of 1637 adolescent girls, special attention is paid to two groups aged at less than or equal to 15 years and 16 to 19 years. The random effects model yielded a *I*^2^of 82% with a pooled OR of good MHM practices between two age groups of 0.77 (95% CI = [0.44, 1.34]), which is not statistically significant (Figure 4a).

**Figure 4a.**
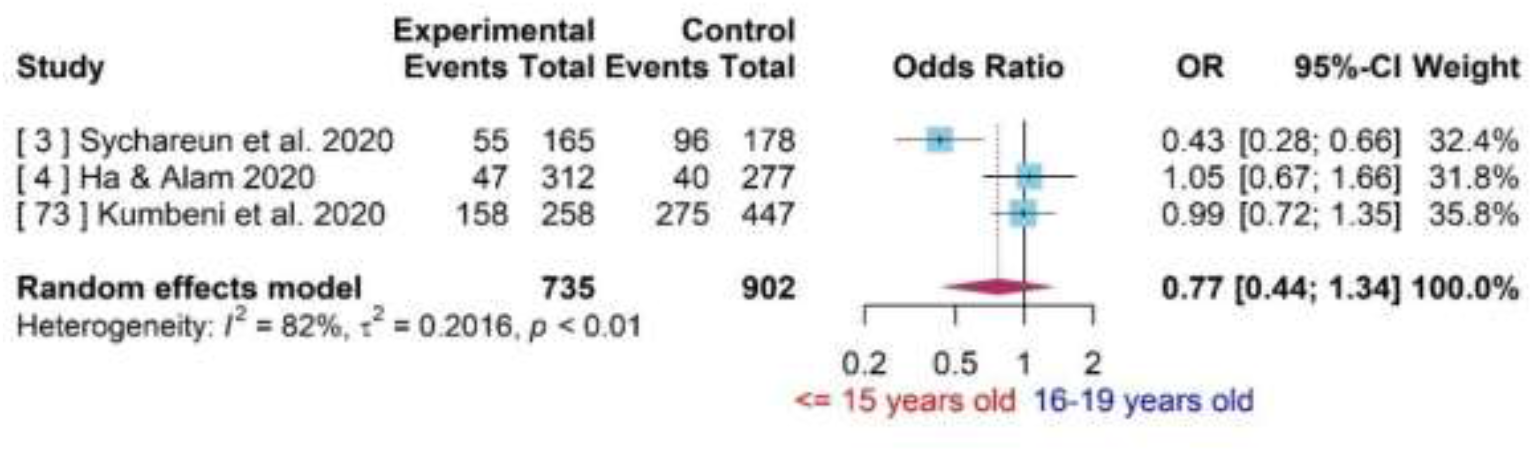
Forest plot for the difference of MHM practice level (Good/Bad) between two age groups.

#### Difference of MHM Practice Level (Good/Bad) among Adolescent Girls with Uneducated and Educated Father/Mother

Parents’ educational background showed an impact on MHM practices among adolescent girls. In some studies father’s or mother’s educational status was divided into illiterate and literate, while in other studies categorised as uneducated, primary education, secondary or high school education, and college or above. To simplify the data, father’s or mother’s educational status was defined as uneducated, where either parent lacked primary education whilst, educated was anyone that had any above secondary. Figure 4b showed a forest plot for difference of MHM practice level (good/bad) among adolescent girls with uneducated and educated fathers, and the pooled OR was 0.55 (95% CI = [0.36,0.83]). This provides significant evidence of the lower prevalence of good MHM practice among adolescent girls with uneducated fathers. Similarly, Figure 4c demonstrated that adolescent girls with uneducated mothers were 0.48 times less likely to have good MHM practices than those with educated mothers.

**Figure 4b.**
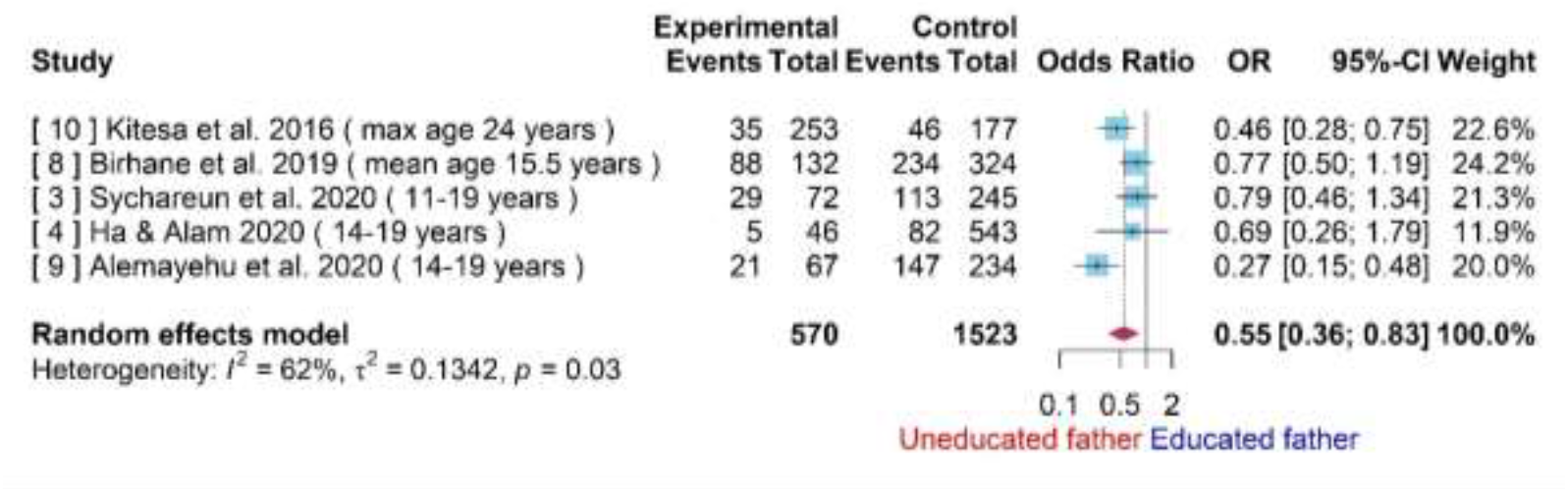
Forest plot for difference of MHM practice level (good/bad) among adolescent girls with uneducated and educated father.

**Figure 4c.**
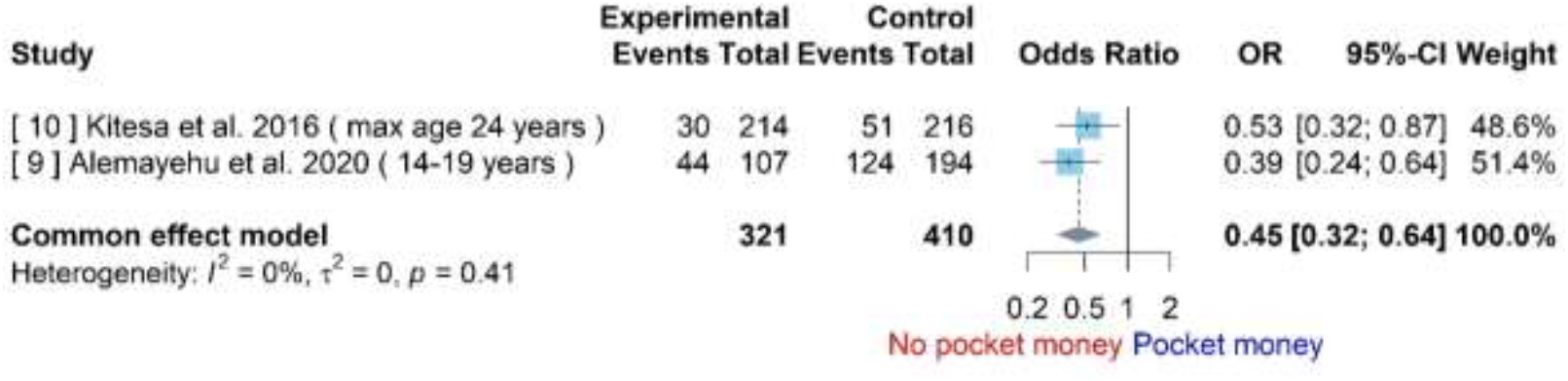
Forest plot for difference of MHM practice level (good/bad) among adolescent girls with uneducated and educated mother. Available in supplemental material

#### Difference of MHM Practice Level (Good/Bad) among Adolescent Girls without and with Pocket Money

To some extent, the possibility of getting a financial allowance, also referred to as a pocket money indicated the socioeconomic status of the family which would indicate their affordability to disposable sanitary pads. Thus, two studies that reported on the use of disposable sanitary pads with a total of 731 adolescent girls were analysed. The forest plot (Figure 5a) demonstrates the difference of MHM practice levels among adolescent girls with and without pocket money. The *I*^2^ = 0 (p-value = 0.41) means that there was very weak statistical heterogeneity. The pooled OR was 0.45 (95%CI = [0.32, 0.64]), indicating that adolescent girls who have no pocket money were 0.55 times less likely to have good MHM practices than those who have pocket money.

**Figure 5a.**
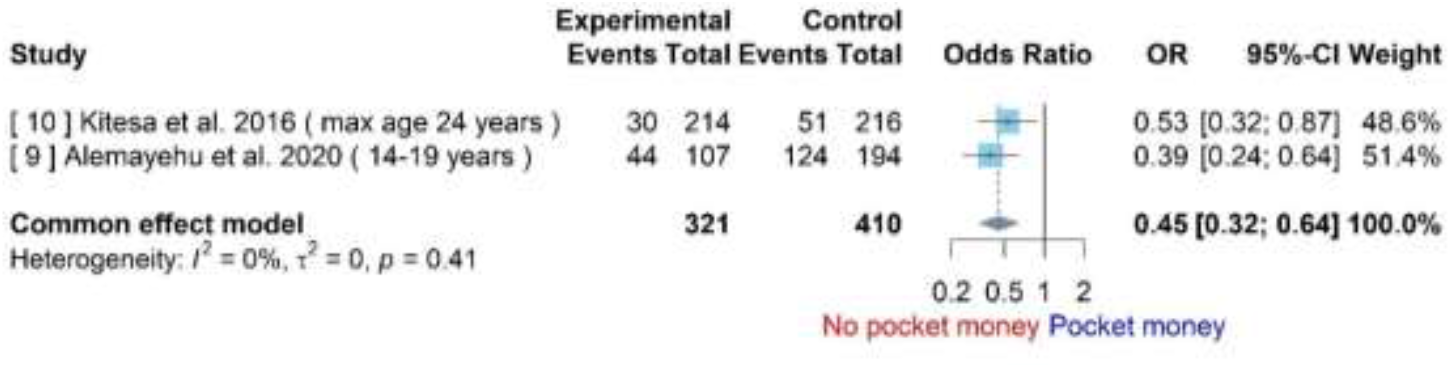
Forest plot for difference of MHM practice level (good/bad) among adolescent girls without and with pocket money.

#### Difference of MHM Practice Level (Good/Bad) among Adolescent Girls of Having No and Having Discussion about Menstruation with Parents

Discussion points between a parent and a young girl were explored where the paradigm indicated open discussions around menstruation issues. Responses to these questions reflects the parent-child relationship. The forest plot indicates (Figure 5b) a difference between MHM practices among adolescent girls who did not have a discussion with their parents versus those who had a discussion was 0.46 (95% CI = [0.28, 0.75]).

**Figure 5b.**
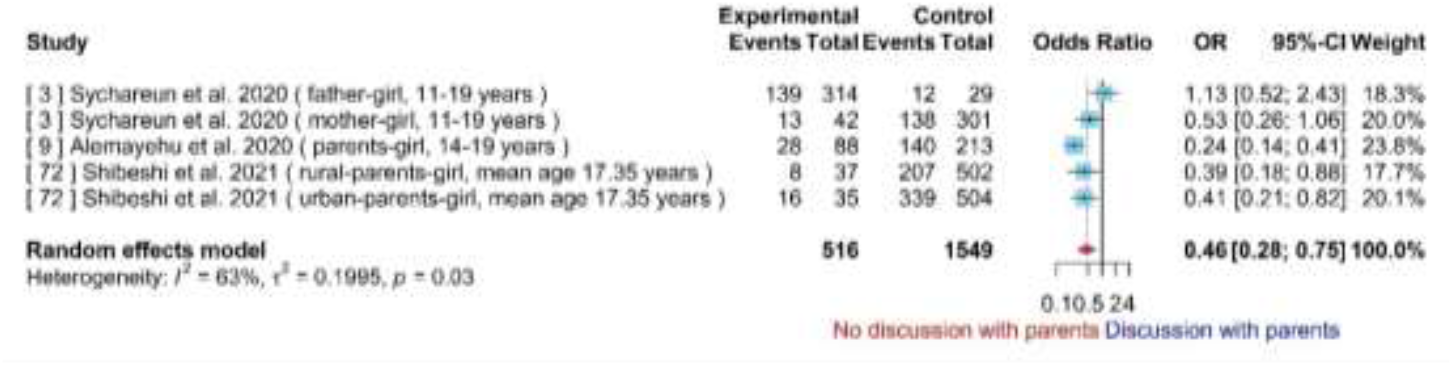
Forest plot for difference of MHM practice level (good/bad) among adolescent girls of having no and having discussion about menstruation with parents. Available in supplemental material

#### Difference of MHM Practice Level (Good/Bad) among Female Followers of Different Religions

Literature indicated the presence of a correlation between religious views and MHM practices, as demonstrated within 3 studies conducted in Ethiopia, with a combined sample size of 1128 adolescent girls. Thus, a pairwise meta-analysis was employed to compare the MHM practice levels among women of Orthodox, Protestant and Islamic beliefs. Figures 5c-5e demonstrated forest plots comparing Orthodox versus Protestant, Protestant versus Islam, and Orthodox versus Islam, respectively. The corresponding pooled ORs were 1.81 (95% CI = [0.47, 6.99]), 0.66 (95% CI = [0.23, 1.92]), and 0.66 (95% CI = [0.87, 1.72]). Based on the CIs, there is no statistically significant difference in MHM practice levels among women from Orthodox, Protestant and Islamic beliefs.

**Figure 5c.**
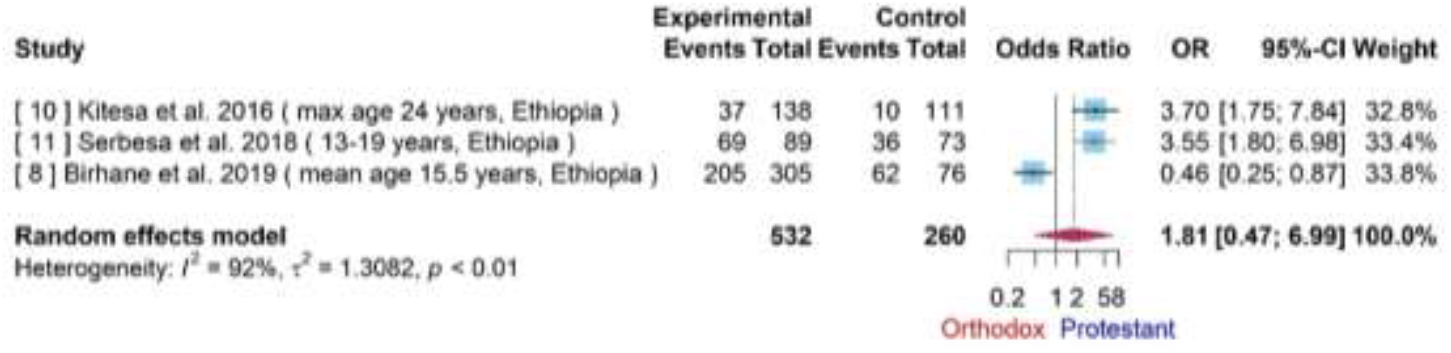
Forest plot for the difference of MHM practice level (Good/Bad) among adolescent girls with orthodox and protestant.

**Figure 5d.**
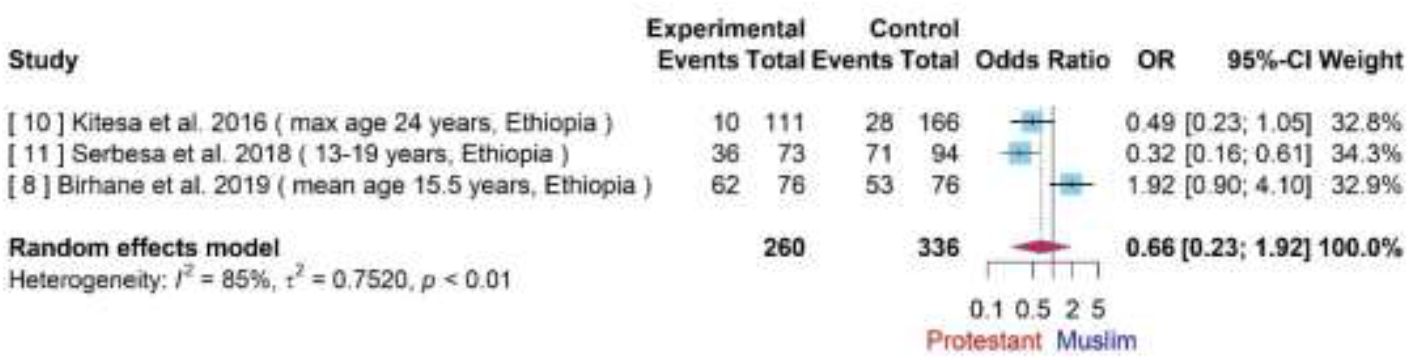
Forest plot for difference of MHM practice level (Good/Bad) among adolescent girls with protestant and Muslim. Available in supplemental material

**Figure 5e.**
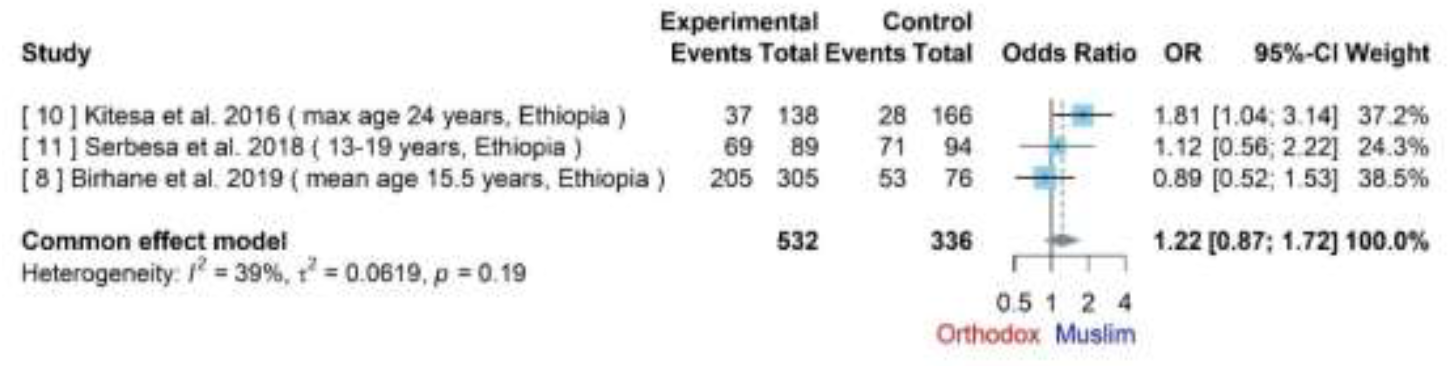
Forest plot for difference of MHM practice level (Good/Bad) among adolescent girls with orthodox and Muslim. Available in supplemental material

#### School Absenteeism Due to Dysmenorrhea

Dysmenorrhea is another key feature of menstruation indicating an important reason for school absenteeism among adolescent girls. Two studies reported mild and moderate menstrual pain among their participants. We combined mild to moderate pain and defined as not severe menstrual pain for the analyses. This was combined with four studies. The total sample size was 1582 (See Supplementary Figure 1). The pooled OR of school absenteeism between adolescent girls with severe and not severe menstrual pain was 4.26 (95% CI = [2.27, 7.99]), indicating those with severe menstrual pain were 4.26 times more likely to miss school than those without menstrual pain.

Participants in study 7 were aged between 19 to 25 years of age, whilst others were less than 19 years old. There appears to be high heterogeneity (*I*^2^=83%, p-value < 0.01) in the sample. Thus, it was excluded, and the heterogeneity was re-evaluated where the pooled OR is 2.98 (95% CI = [2.29, 3.87]). Supplementary Figure 2 indicates *I*^2^ to be 0 with a p-value of < 0.01. The heterogeneity, therefore, was specific to Study 7. The participant group of 19 to 25 years old, or, more precisely, age group may be one of the main sources of heterogeneity.

To explore the association between school absenteeism and whether or not using disposable sanitary pads have any impact, a meta-analysis was applied to 3 studies with a total sample size of 1280 adolescent girls. Supplementary Figure 3 indicated significant evidence of statistical heterogeneity (*I*^2^=83%, p-value < 0.01). The pooled OR of 2.08 (95% CI = [1.10, 3.91]) indicates that adolescent schoolgirls who did not use disposable sanitary pads were 1.08 times more likely to be absent from school than those using sanitary pads.

#### Association between Dysmenorrhea and Regularity of Menstrual Cycle

As presented in the former part, there is a statistically significant association between the severity of dysmenorrhea and school absenteeism. To further identify the possible causes of dysmenorrhea, two studies were meta-analysed with a total sample size of 1285 with confirmed experience of regular or irregular menstrual cycles. Supplementary Figure 4 indicated a lack of statistical heterogeneity (*I*^2^ = 0, p-value = 0.89) and thus the fixed effects model was used. The pooled OR was 2.31 (95% CI = [1.76,3.02]), indicating the prevalence of dysmenorrhea among adolescent girls with irregular menstrual cycles is 2.31 times as high as those with a regular cycle.

Another key area of period poverty is the associated mental health impact, which can differ between those who suffer from mental illness and those who do not. Whilst there was insufficient data for a meta-analysis, there was evidence to suggest a link between menstruation and prevalence of stress, anxiety, and depression. ^3^ In addition, socioeconomic status can impact the prevalence of stress, anxiety and depression experienced by different populations.

## Discussion

Period poverty is a global health issue, more prominent in low-middle-income countries. There are varying risk factors dependent on geographical location and this reflects in the differing sociological and clinical features, as explored in this paper.

This study demonstrates correlations between severity of dysmenorrhea and school absenteeism among girls between 14-19 years of age with and without regular menstruation. Another key area of period poverty is the associated mental health impact, which can differ between those who suffer from mental illness and those who do not. Whilst there was insufficient data for a meta-analysis, there was evidence to suggest a link between menstruation and prevalence of stress, anxiety, and depression. ^3^ In addition, socioeconomic status can impact the prevalence of stress, anxiety and depression experienced by different populations. This could be exacerbated among those acquiring urinary tract infection (UTIs). ^35-36^

UTIs have been reported by Das and colleagues to be a common problem among those without pad use. Janoowalla and colleagues demonstrated no change in the prevalence of urinary tract infections between those using and not-using pads in the Kibogora region in Rwanda. Das and colleagues indicated a higher risk of urogenital infections among women using reusable absorbent pads within the Odisha region in India. Bacterial vaginosis (BV) is another issue impacting women with poorer menstrual practices. Das and colleagues reported that menstrual hygiene practices were associated with a symptomatic BV or UTI. ^36^

MHM practices were another key endpoint in this study which demonstrated to differ among women of Islamic, Protestant and Orthodox religious beliefs in Ethiopia. Representativeness of these findings to other ethnicities requires further research.

Homelessness is another facet that is vital to explore to identify the impact of period poverty. For example, in the United States, 553,000 experience homelessness in a night compared to 320,00 in the UK. ^26-27^ It is reported that 25% of homeless service users in the UK are single women and 28% in the United States. These could be underestimated as hidden homeless is another facet where people do not access services but stay in temporary accommodation settings, including friends and relatives. Many official reports lack information regarding experiences of menstruation among the homeless. Padgett and colleagues demonstrate that this multifaceted concern or as a feature of reproductive health is now being explored, although comprehensive evidence is required. ^28^ Phenomenology demonstrates homeless menstrual lived experiences, which is an important aspect of understanding period poverty by exploring the interrelatedness between the consciousness, body and the flesh of a woman. ^29^ Homeless people are a marginalised community; thus, menstruation could emphasise their vulnerability. Historically, social sciences research has focussed on commodification, medicalisation and stigma associated with menstruation. Anglo-American publications have also failed to discuss intersectionality and focus primarily on white, middle-class, cisgender women or in a developmental context where women are in poverty. ^30-32^ The sociopsychological and socioeconomic aspects associated with women living in poverty versus those not can sometimes be polarised from a period poverty perspective. Health outcomes among disenfranchised groups of women due to lack of or minimal access to menstrual products can have significant effects where clinical interventions would be required to manage the symptoms, including systemic issues. Another facet is that the supply of menstrual products to homeless services such as shelters, and day centres should be more effective. This should include the availability of staff that could be approached to talk about menstruation or any associated problems. ^33^

Another facet of period poverty is the composition of menstrual pads which is not the same in terms of their textile and polymer composition. Varying viscosity of menstrual flow could also impact the suitability of the differing pads, given that these are worn for different periods. Another aspect to consider as an external factor for menstrual products would be to produce material that can be disposed of in a biodegradable manner. Mazgak and colleagues (2006) and Hait et al. (2019) indicated that sanitary pads have a higher negative environmental footprint due to eutrophication and climate change. Limited evidence is available about menstrual underwear and menstrual cups associated with environmental impact. This further complicates menstrual hygiene issues, equitable availability and acceptability, especially among LMIC populations.

Whilst this study has indicated the majority of the evidence on period poverty is within LMIC and MICs, there appears to be a lack of studies available within developed countries despite the definition of “period poverty”, including the inability to afford menstrual products. Given the risk of living costs, many media publications and social media posts indicate that period poverty is a concern within developed countries. For example, Cardoso and colleagues indicated that women in the United States reported 14.2% experienced period poverty in 2020, with an additional 10% experiencing it monthly. ^36^ Whilst knowledge, attitude and practices associated with menstruation among poorer and vulnerable communities are likely to be lower regardless of the geographical location. As a result, the psychosocial dynamics may have a negative impact. The findings of these studies may have been exacerbated due to the COVID-19 pandemic with substantial increases in unemployment and cost of living. Basic goods and service cost increase includes those of menstrual products. Thus, the pandemic has had gendered implications impacting the vulnerability of women. Caretaker roles of women have significantly grown as a result of lockdowns, and such requirements have been inadequately explored. ^42^

In addition, limited evidence is demonstrated around mental health implications due to the period of poverty. Cardoso and colleagues demonstrated an association between period poverty and depression among women within the United States who were previously depression naïve.^36^ This is similar to the findings reported between food insecurity associated with depression in adults and anxiety, depression and suicidal ideation among adolescents. ^36-39^ Similarly, depression and anxiety were reported among people experiencing housing insecurities compared to those with stable houses. ^40-41^

### Limitations

The data identified is limited to either smaller sample sizes and/or geographical locations that could limit the generalisability of the findings to introduce impactful and meaningful changes to policy and clinical practice.

## Conclusion

Undoubtedly, many issues affect the experiences of managing menstruation and access to menstrual products. Thus, there could be a growing need for global period poverty. To address the current gaps and ensure period poverty can be minimised, comprehensive research would be required. Policymakers and independent authorities should consider improved healthcare legislation, equitable access to menstrual products, information, and healthcare providers.

## Data Availability

All data produced in the present study are available upon reasonable request to the authors

## Abbreviations

(MHM): Menstrual hygiene management
(WHO): World Health Organisation
(UNICEF): United Nations International Children’s Emergency Fund
(LMICs): Low-middle-income countries
(MICs): Middle-low-income countries
(HICs): High-income countries

## Acknowledgements

This study has been supported by Southern Health NHS Foundation Trust.

## Supplementary Figures

**Supplementary Figure 1.**
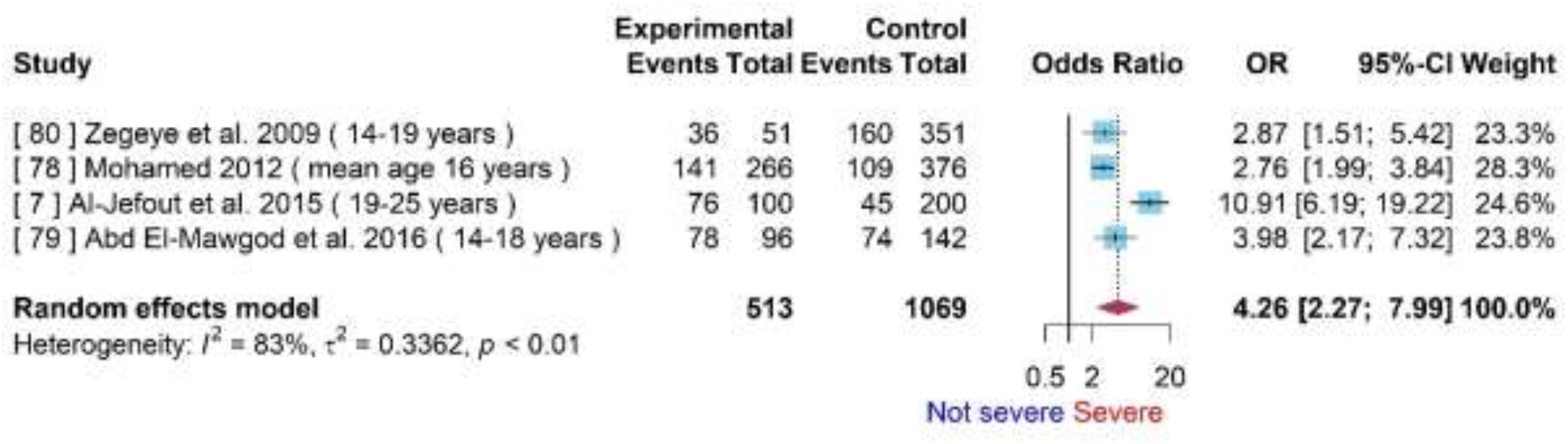
Based on four studies, the forest plot for the association between school absenteeism and severity of menstrual pain.

**Supplementary Figure 2.**
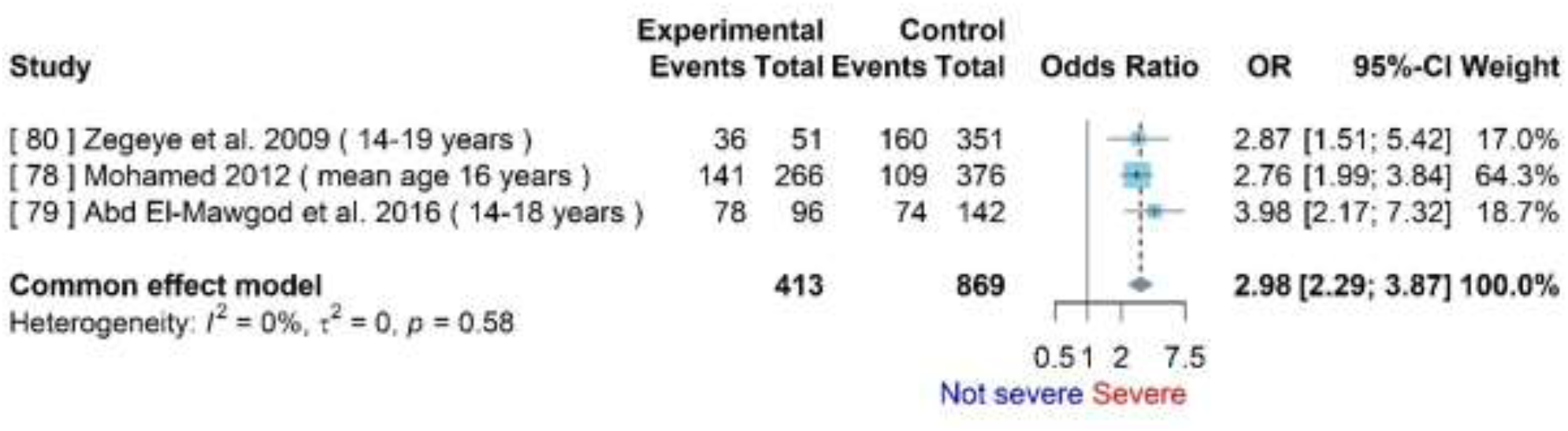
Forest plot for the association between school absenteeism and severity of menstrual pain with study 7 removed.

**Supplementary Figure 3.**
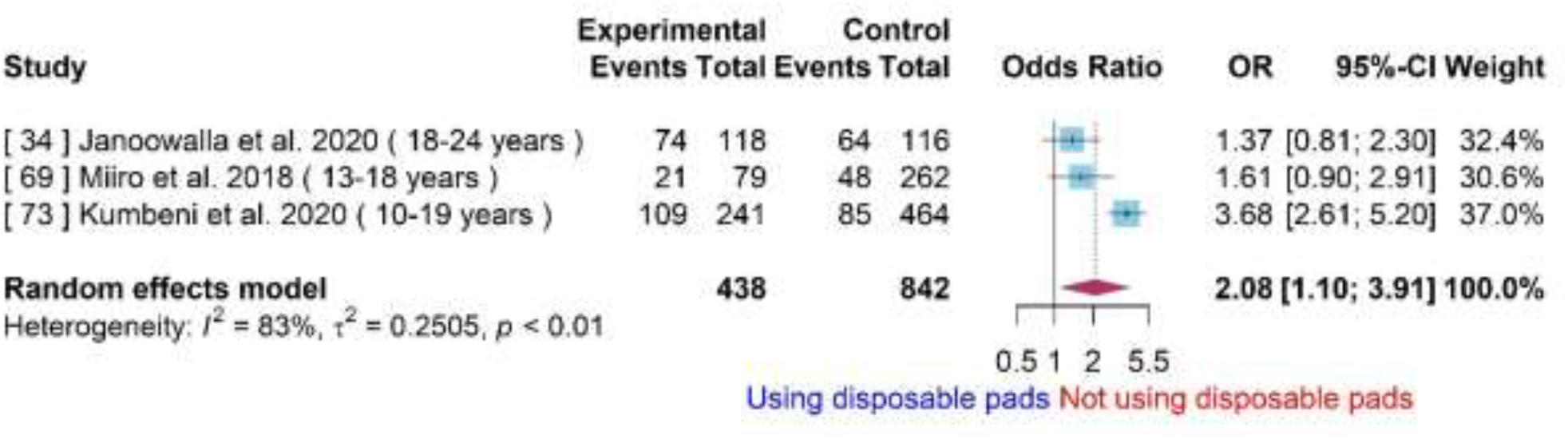
Forest plot for the association between school absenteeism and use of sanitary pads during menstruation.

**Supplementary Figure 4.**
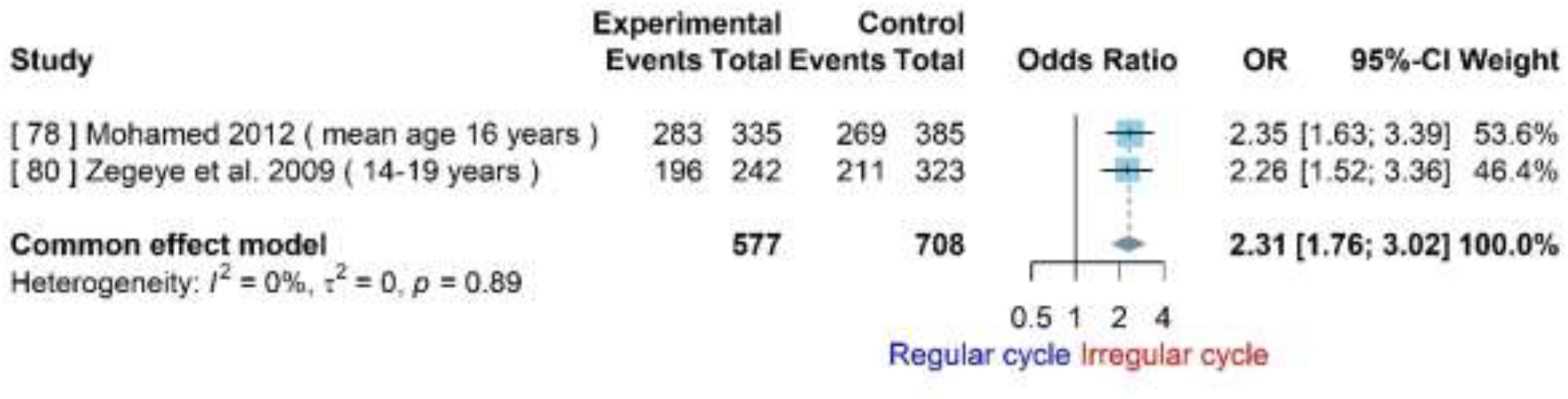
Forest plot for the association between dysmenorrhea and regularity of menstrual cycle.

